# Phenotyping Refractory Cardiogenic Shock Patients Receiving Venous-arterial Extracorporeal Membrane Oxygenation with Machine Learning Algorithms

**DOI:** 10.1101/2023.04.20.23288900

**Authors:** Shuo Wang, Liangshan Wang, Zhongtao Du, Feng Yang, Xing Hao, Xiaomeng Wang, Chengcheng Shao, Chenglong Li, Hong Wang, Xiaotong Hou

**Affiliations:** Center for Cardiac Intensive Care, Beijing Anzhen Hospital, Capital Medical University, Beijing 100029, P.R. China.

**Author notes:** Shuo Wang and Liangshan Wang contributed equally to this paper. Correspondence to: **Xiaotong Hou, MD, PhD** Center for Cardiac Intensive Care, Beijing Anzhen Hospital, Capital Medical University, 2 Anzhen Rd, Chaoyang District, Beijing 100029, P.R. China.; E-mail address, **Hong Wang, MD, PhD** Center for Cardiac Intensive Care, Beijing Anzhen Hospital, Capital Medical University, 2 Anzhen Rd, Chaoyang District, Beijing 100029, P.R. China.; E-mail address, **Chenglong Li, MD** Center for Cardiac Intensive Care, Beijing Anzhen Hospital, Capital Medical University, 2 Anzhen Rd, Chaoyang District, Beijing 100029, P.R. China.

**Keywords:** Extracorporeal membrane oxygenation, cardiogenic shock, phenotypes, clusters, machine learning

## Abstract

**Background:** Refractory cardiogenic shock (CS) patients receiving venous-arterial extracorporeal membrane oxygenation (VA-ECMO) have a wide range of mortality, machine algorithm methods may explain the potential heterogeneity of these patients.

**Methods:** Between January 2018 and May 2021, 210 patients with CS who were receiving VA-ECMO support were enrolled and analyzed retrospectively. The k-means consensus agnostic algorithm was used. Patients were divided into three clusters based on covariates, such as platelet count (PLT), aspartic acid transaminase (AST), Interleukin-6 (IL-6), prothrombin time (PT), and serum lactate level 24 hours after ECMO initiation. The clinical and laboratory profiles were analyzed.

**Results:** Among 210 CS with CS receiving ECMO, 148 (70.5%) were men, with a median age of 62 years (interquartile range (IQR): 53-67). Overall, 104 (49.5%) patients survived to discharge with 142 (67.6%) survived on ECMO. The patients were phenotyped into three clusters: (1) “platelet preserved (I)” Phenotype (36 [17.1%] patients), characterized by a preserved platelet count; (2) “hyperinflammatory (II)” phenotype (72 [34.3%] patients), characterized by a significant inflammatory response with higher Interleukin-6 (IL-6), and Interleukin-10 (IL-10) levels; and (3) “hepatic-renal (III)” phenotype (102 [48.6%] patients), characterized by unfavorable conditions in creatinine, aspartic acid transaminase, alanine aminotransferase, direct bilirubin, and prothrombin time. The in-hospital mortality rates were 25.0%, 52.8%, and 55.9% for phenotypes I, II, and III, respectively (P = 0.005).

**Conclusion:** The consensus k-means algorithm analysis identified three phenotypes in refractory patients with CS receiving VA-ECMO: “platelet preserved,” “hyperinflammatory,” and “hepatic-renal.” The phenotypes are associated with the clinical profile and mortality, allowing treatment strategies for subsets of patients with CS receiving ECMO to be developed.

## Introduction

Cardiogenic shock (CS) is associated with a mortality rate of up to 50% [1]. Venoarterial extracorporeal membrane oxygenation (VA-ECMO) is a temporary mechanical circulatory support device used to treat CS. Despite the lack of randomized clinical trials confirming the efficacy of VA-ECMO, its use during refractory CS management has increased in recent decades, with a survival rate ranging from 16%–42% [2–4].

CS is a heterogeneous clinical condition. Patients may experience CS as a result of the slow progression of chronic heart failure or after an acute cardiac insult such as acute myocardial infarction (AMI), myocarditis, malignant ventricular arrhythmia, or even cardiac arrest. Thus, patients receiving VA-ECMO have various courses and mortality rates depending on the etiology of CS. The complexity of clinical profiles after ECMO also leads to heterogeneity [5]. These outcomes have been linked to lactate behavior, platelet count, organ function, and inflammation [6–10]. Heterogeneity makes clinical practice more difficult and limits our ability to develop new strategies in unselected populations.

ECMO scoring systems such as the survival after veno-arterial-ECMO (SAVE), prediction of cardiogenic shock outcome for acute myocardial infarction(AMI) patients salvaged by VA-ECMO (ENCOURAGE), and predicting mortality in patients undergoing veno-arterial extracorporeal membrane oxygenation after coronary artery bypass grafting (REMEMBER) scores, have been developed, to predict prognosis and potentially identify patients who may benefit from ECMO using pre-ECMO parameters [3, 10, 11]. However, they were unable to characterize patients on ECMO. Therefore, determining and understanding the heterogeneity of the illness status beyond CS causes and pre-ECMO parameters may help clinicians to distinguish phenotypes and translate them into new therapies.

Machine learning (ML) approaches focus on categorizing certain data points into single distinguished datasets, which have been previously implemented to define heterogeneous clinical syndromes such as acute respiratory distress syndrome (ARDS), sepsis, trauma, acute kidney injury, and CS [12–14]. Herein, we designed the ML method to explore the potential heterogeneity of patients with CS receiving VA-ECMO and review the multiple dimensions of these patients, including clinical and biological features, as well as inflammatory status after VA ECMO initiation and phenotype of these patients into certain single subgroups (clusters). This could further our understanding of the physiology of CS patients with ECMO, select subgroups of patients for potential interventions in clinical trials, and be incorporated into clinical practice as a risk assessment tool.

## Method

### Patient population

This study was a single-center, prospective, observational study approved by the institutional ethics review board (IRB) at Beijing Anzhen Hospital (202102X). International Research Database for Extracorporeal Support (approval date: February 23, 2021; study title: International Research Database for Extracorporeal Support). All the procedures were followed in accordance with the ethical standards of the responsible committee on human experimentation and with the Helsinki Declaration of 1975. Informed consent for demographic, physiological and hospital-outcome data analyses was not obtained because this observational study did not modify existing diagnostic or therapeutic strategies. However, patients and/or relatives were informed about the anonymous data collection and that they could decline inclusion.

The study enrolled adult patients with CS who were receiving VA-ECMO for circulatory support. CS is defined as follows [15]: (1) systolic blood pressure < 90 mmHg for 30 min, mean arterial pressure < 65 mmHg for 30 min, or the need for vasopressors to achieve a blood pressure of 90 mmHg; (2) pulmonary congestion or elevated left ventricular filling pressure; and (3) signs of impaired organ perfusion with at least one of the following indications: altered mental status, cold, clammy skin, oliguria, or increased serum lactate despite optimized supportive measures, such as an intra-aortic balloon pump and inotropes.

Between January 2018 and May 2021, 282 patients receiving ECMO at the Beijing Anzhen Hospital were screened, with 210 patients eventually recruited and analyzed for this study. Seventy-two patients were excluded for the following reasons: age < 18 years (17), acute respiratory failure treated with venovenous ECMO (4), VA-ECMO duration < 24 hours (6), severe missing clinical data (7), and failure to obtain informed consent were excluded (6) (**Figure 1A**).

**Figure 1.**
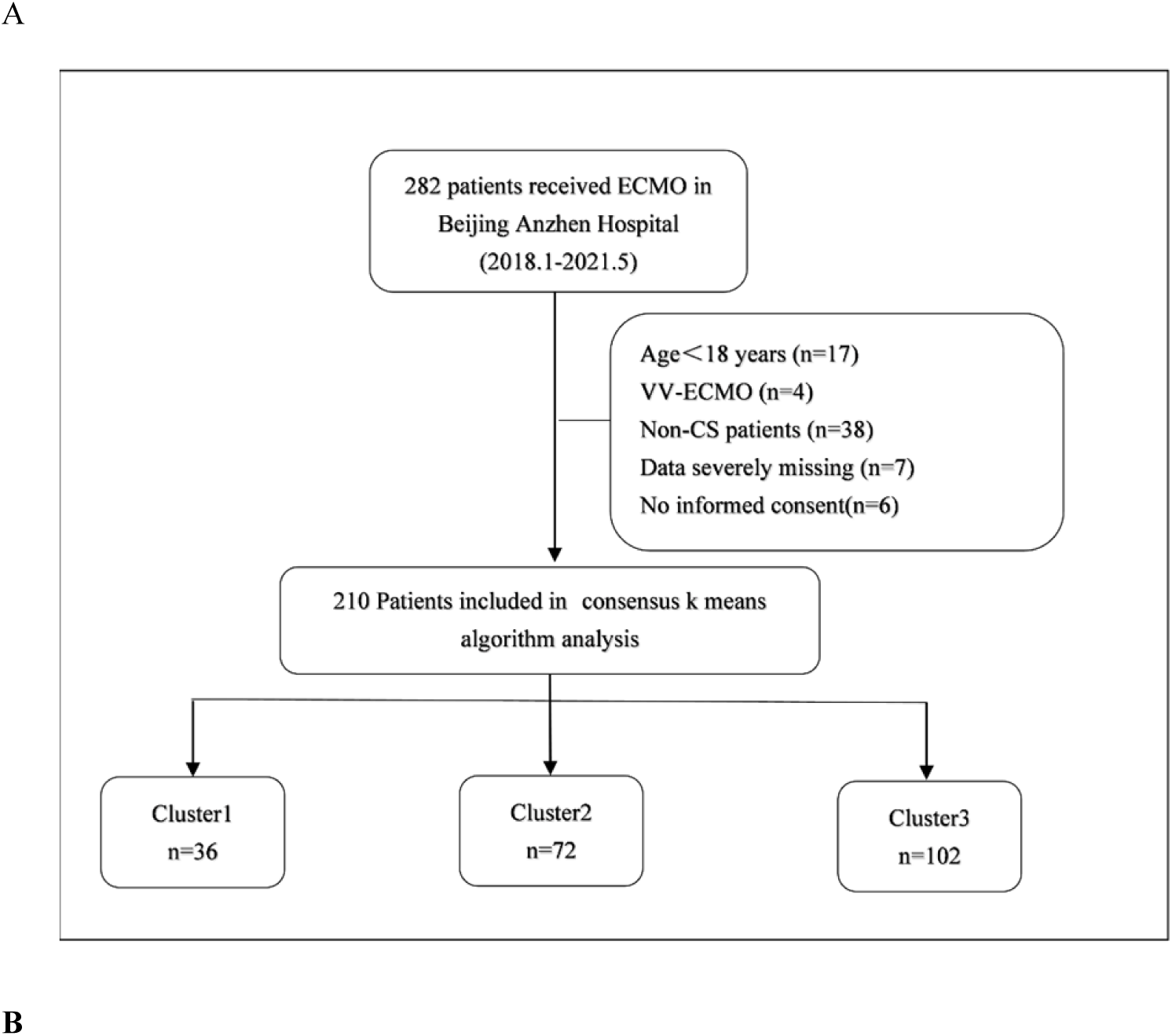
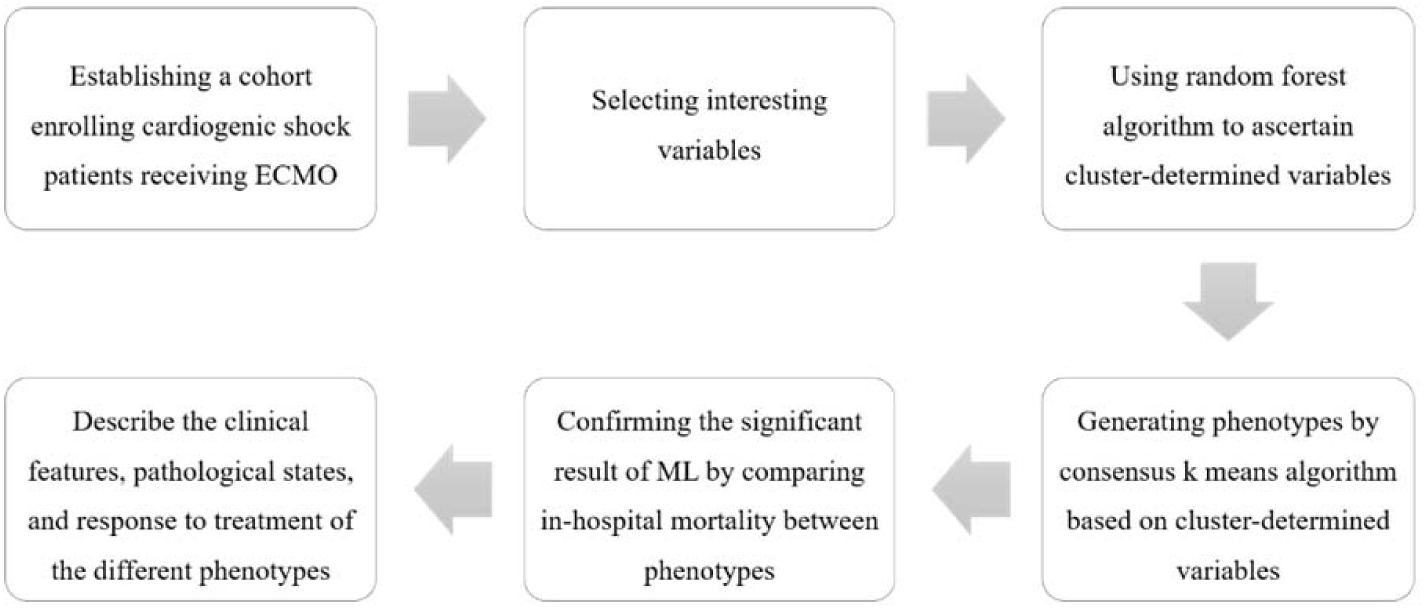
Flow diagram for patient selection (**A**) and machine learning algorithms (**B**).

### ECMO management

Details regarding VA-ECMO initiation and management have been previously described [16]. All VA-ECMO procedures were performed by the ECMO team members. VA-ECMO support was initiated via peripheral cannulation via the femoral route, using semi-open or percutaneous methods. An additional 6 Fr catheter was inserted distally into the femoral artery to prevent severe leg ischemia. Clinical assessments were used to adjust ECMO blood flow (e.g., mixed venous oxygen saturation, evidence of hypoperfusion, resolution of hyperlactatemia, and normalization of mean arterial pressure). Unfractionated heparin was administered intravenously to maintain an activated clotting time of 180–210 s or an activated partial thromboplastin time of 1.5–2 times normal. The complications associated with ECMO were closely monitored. Patients who met our published institutional weaning criteria and passed an ECMO weaning trial consisting of decreasing and clamping the ECMO flow were given ECMO weaning [16]. Successful ECMO weaning was defined as the lack of obvious hemodynamic deterioration for at least 48 h after the removal of ECMO support.

### Selection of cluster-determined variables

Baseline characteristics were recorded within the initial 24 h after ICU admission, including age, sex, body mass index (BMI), laboratory test after 24 h of ECMO initiation (including complete blood count, hepatic-renal, and coagulation function), and arterial blood gas (before ECMO initiation, 4 h, and 24 h after ECMO initiation). As the inflammatory response is generally regarded as a crucial phenomenon relevant to ECMO patient outcomes [10, 19–22], interleukin-6 (IL-6) and interleukin-10 (IL-10) were detected. Moreover, treatment details such as body temperature during ECMO, ECMO peak flow, pre-ECMO left ventricular ejection fraction (LVEF), left ventricular diastolic diameter, and mean arterial pressure were also considered. The use of vasoactive agents was described as the vasoactive inotropic score (VIS), which was calculated when the vasoactive agents adequately maintained a relatively stable hemodynamic status according to the following formula: dosages of dopamine (in µg.kg^-1^.min^-1^) + dosages of dobutamine (in µg.kg^-1^.min^-1^) + [dosages of epinephrine (in µg.kg^-1^.min^-1^) + norepinephrine (in µg.kg^-1^.min^-1^)] × 100 + dosages of pituitrin(in unit/min) * 100 +dosages of milrinone (in µg.kg^-1^.min^-1^) * 15 [19].

The candidate variables used as class-defining parameters in the model were selected according to previous research, which had a specific association with patient outcomes [6, 8, 23]. Some indicators that may be related to patient outcomes according to clinical experience were also taken into consideration. A previous study recommended that the minimal sample size of cluster analysis was more than 2^n cases (n = number of variables), and 5 × 2^n would be favorable [24]. Based on this, a random forest classifier was used to identify important variables according to mortality association before applying the clustering algorithm. Because the random forest classifier could not identify colinear variables, we first ran the random forest classifier with all variables of interest to ascertain their predictive value for in-hospital mortality. We then trained the random forest classifier again after the removal of correlating variables and identified the most predictive variables for the subsequent clustering process. The identified variables with the highest predictive value were then selected and deemed as cluster-determined variables, which were further used to define the optimal number of clusters (k).

### Consensus k-means algorithm analysis

Consensus k-means algorithm analysis is a classic ML technique performed as a method in previously reported research to identify the homogeneity of a specific disease. Cluster-determined covariates were selected before performing the cluster analysis (as mentioned above). Before starting the consensus k-means analysis, the number of clusters (k) was confirmed using the random forest classifier and several other methods (cumulative distribution function plot, cluster-consensus plot, elbow plot, and t-distributed stochastic neighbor embedding (TSNE) plot). In the present study. A k-value of 3 was considered favorable. **Figure 1B** shows a flowchart of the ML algorithms. All the main machine learning steps were carried out using R-software on RStudio 2021.09.1, building 372.

### Statistics analyses

Continuous variables were compared using the Student’s t-test or Wilcoxon’s rank test, while categorical variables were compared using the chi-square test or Fisher’s exact test. Continuous variables were normalized and presented as median (interquartile range) and compared using chi-square tests or Fisher’s exact tests, while categorical variables were presented as numbers (percentages) and compared using Wilcoxon’s signed-rank tests. The superscripts a, b, and c indicate significant pairwise differences among the clusters. Statistical analyses were performed using Statistical Package for the Social Sciences (SPSS) (version 25.0, IBM, New York, USA) and R-software on RStudio 2021.09.1 build 372. The data were visualized using R-software on RStudio 2021.09.1 build 372 and Prism 8 Version 8.0.2(263). Statistical significance was set at a two-tailed p-value of < 0.05.

## Result

### Patient characteristics

Between January 2018 and May 2021, 210 patients with CS receiving VA-ECMO were enrolled for the final analysis in the present study (**Figure 1A**). The median patient age was 62 years (IQR: 53-67 years). The study included 148 men (70.5%). The rates of successful weaning from ECMO and in-hospital mortality were 67.6% and 49.5%, respectively. The baseline characteristics of the patients are presented in **Table 1**. The etiology of CS (some patients had multiple diagnoses) included coronary artery disease (109), valvular heart disease (81), congenital heart disease (4), myocarditis (8), and aortic artery dissection (13). One hundred and forty-four patients presented with postcardiotomy CS and received ECMO. The median duration of ECMO was 105.4 h (IQR: 66.7-153.6).

**Table 1.**
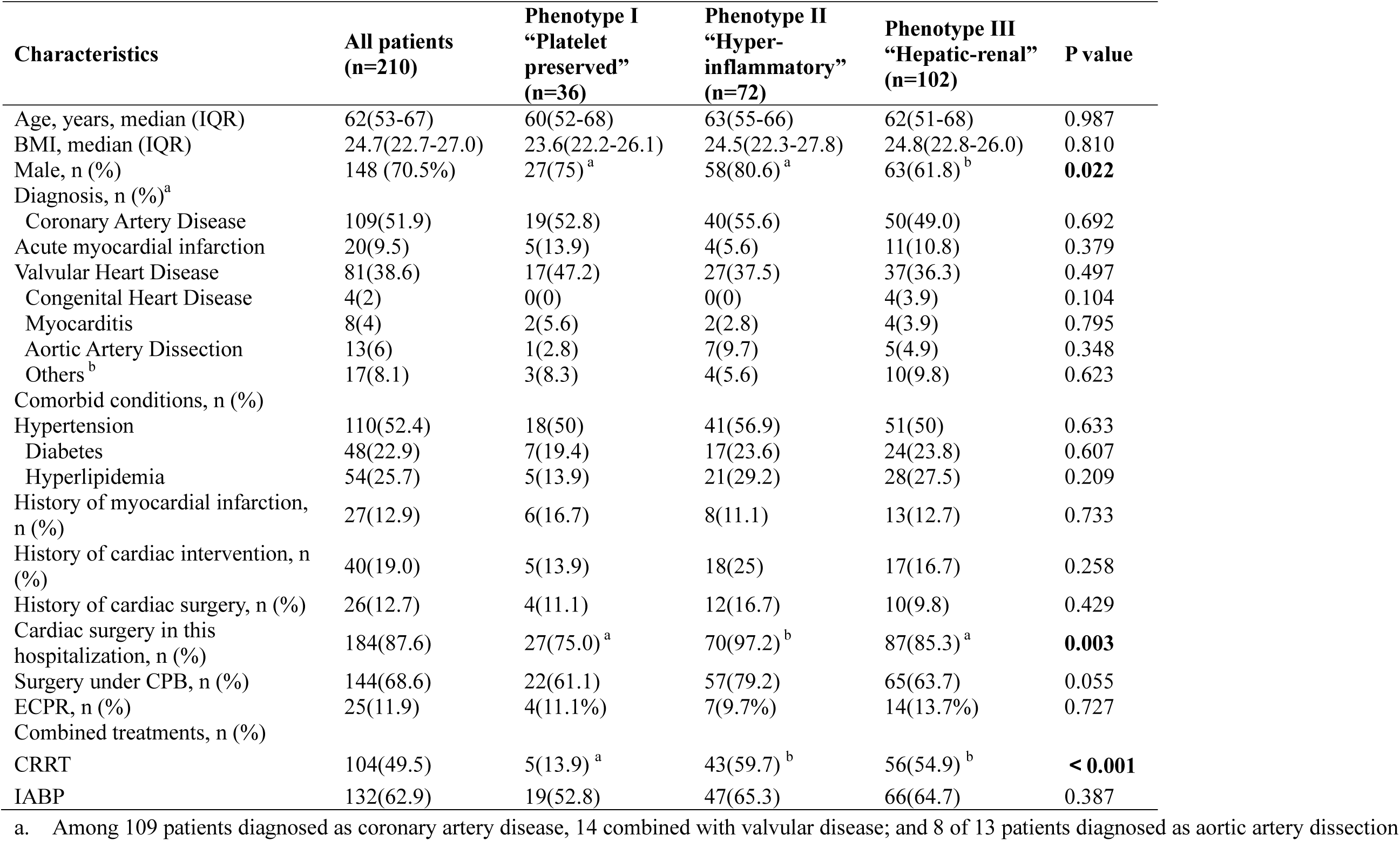
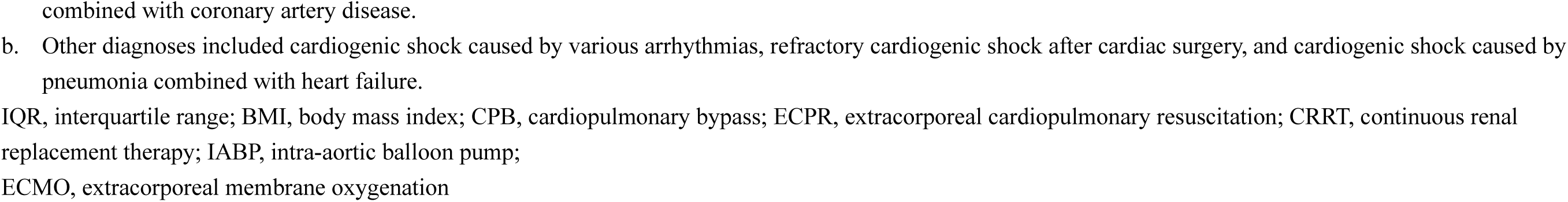
Patient Characteristics

| Characteristics | All patients<br>(n=210) | Phenotype I<br>“Platelet<br>preserved”<br>(n=36) | Phenotype II<br>“Hyper-<br>inflammatory”<br>(n=72) | Phenotype III<br>“Hepatic-renal”<br>(n=102) | P value |
| --- | --- | --- | --- | --- | --- |
| Age, years, median (IQR) | 62(53-67) | 60(52-68) | 63(55-66) | 62(51-68) | 0.987 |
| BMI, median (IQR) | 24.7(22.7-27.0) | 23.6(22.2-26.1) | 24.5(22.3-27.8) | 24.8(22.8-26.0) | 0.810 |
| Male, n (%) | 148 (70.5%) | 27(75)<sup>a</sup> | 58(80.6)<sup>a</sup> | 63(61.8)<sup>b</sup> | **0.022** |
| Diagnosis, n (%)<sup>a</sup> |  |  |  |  |  |
| Coronary Artery Disease | 109(51.9) | 19(52.8) | 40(55.6) | 50(49.0) | 0.692 |
| Acute myocardial infarction | 20(9.5) | 5(13.9) | 4(5.6) | 11(10.8) | 0.379 |
| Valvular Heart Disease | 81(38.6) | 17(47.2) | 27(37.5) | 37(36.3) | 0.497 |
| Congenital Heart Disease | 4(2) | 0(0) | 0(0) | 4(3.9) | 0.104 |
| Myocarditis | 8(4) | 2(5.6) | 2(2.8) | 4(3.9) | 0.795 |
| Aortic Artery Dissection | 13(6) | 1(2.8) | 7(9.7) | 5(4.9) | 0.348 |
| Others<sup>b</sup> | 17(8.1) | 3(8.3) | 4(5.6) | 10(9.8) | 0.623 |
| Comorbid conditions, n (%) |  |  |  |  |  |
| Hypertension | 110(52.4) | 18(50) | 41(56.9) | 51(50) | 0.633 |
| Diabetes | 48(22.9) | 7(19.4) | 17(23.6) | 24(23.8) | 0.607 |
| Hyperlipidemia | 54(25.7) | 5(13.9) | 21(29.2) | 28(27.5) | 0.209 |
| History of myocardial infarction, n (%) | 27(12.9) | 6(16.7) | 8(11.1) | 13(12.7) | 0.733 |
| History of cardiac intervention, n (%) | 40(19.0) | 5(13.9) | 18(25) | 17(16.7) | 0.258 |
| History of cardiac surgery, n (%) | 26(12.7) | 4(11.1) | 12(16.7) | 10(9.8) | 0.429 |
| Cardiac surgery in this hospitalization, n (%) | 184(87.6) | 27(75.0)<sup>a</sup> | 70(97.2)<sup>b</sup> | 87(85.3)<sup>a</sup> | **0.003** |
| Surgery under CPB, n (%) | 144(68.6) | 22(61.1) | 57(79.2) | 65(63.7) | 0.055 |
| ECPR, n (%) | 25(11.9) | 4(11.1%) | 7(9.7%) | 14(13.7%) | 0.727 |
| Combined treatments, n (%) |  |  |  |  |  |
| CRRT | 104(49.5) | 5(13.9)<sup>a</sup> | 43(59.7)<sup>b</sup> | 56(54.9)<sup>b</sup> | **< 0.001** |
| IABP | 132(62.9) | 19(52.8) | 47(65.3) | 66(64.7) | 0.387 |

a. Among 109 patients diagnosed as coronary artery disease, 14 combined with valvular disease; and 8 of 13 patients diagnosed as aortic artery dissection
combined with coronary artery disease.
- b. Other diagnoses included cardiogenic shock caused by various arrhythmias, refractory cardiogenic shock after cardiac surgery, and cardiogenic shock caused by pneumonia combined with heart failure.
IQR, interquartile range; BMI, body mass index; CPB, cardiopulmonary bypass; ECPR, extracorporeal cardiopulmonary resuscitation; CRRT, continuous renal replacement therapy; IABP, intra-aortic balloon pump; ECMO, extracorporeal membrane oxygenation

### Clusters identification

First, a random forest classifier was used to identify variables that predicted in-hospital mortality. Seven significant variables were obtained (AST, 24-hour lactate level, PT, IL-6, ALT, platelet count, and APTT) (**Figure 2A**). As the linear relationships between the seven variables might affect the reliability of the consensus k-means algorithm analysis, a correlation test was performed to eliminate the non-orthogonal variables and reduce the dimensionality of the model. Two variables (APTT and ALT) were excluded (**Figure 2B**). The five highest predictive-value variables were then included in further cluster analysis (**Figure 2C**).

**Figure 2.**
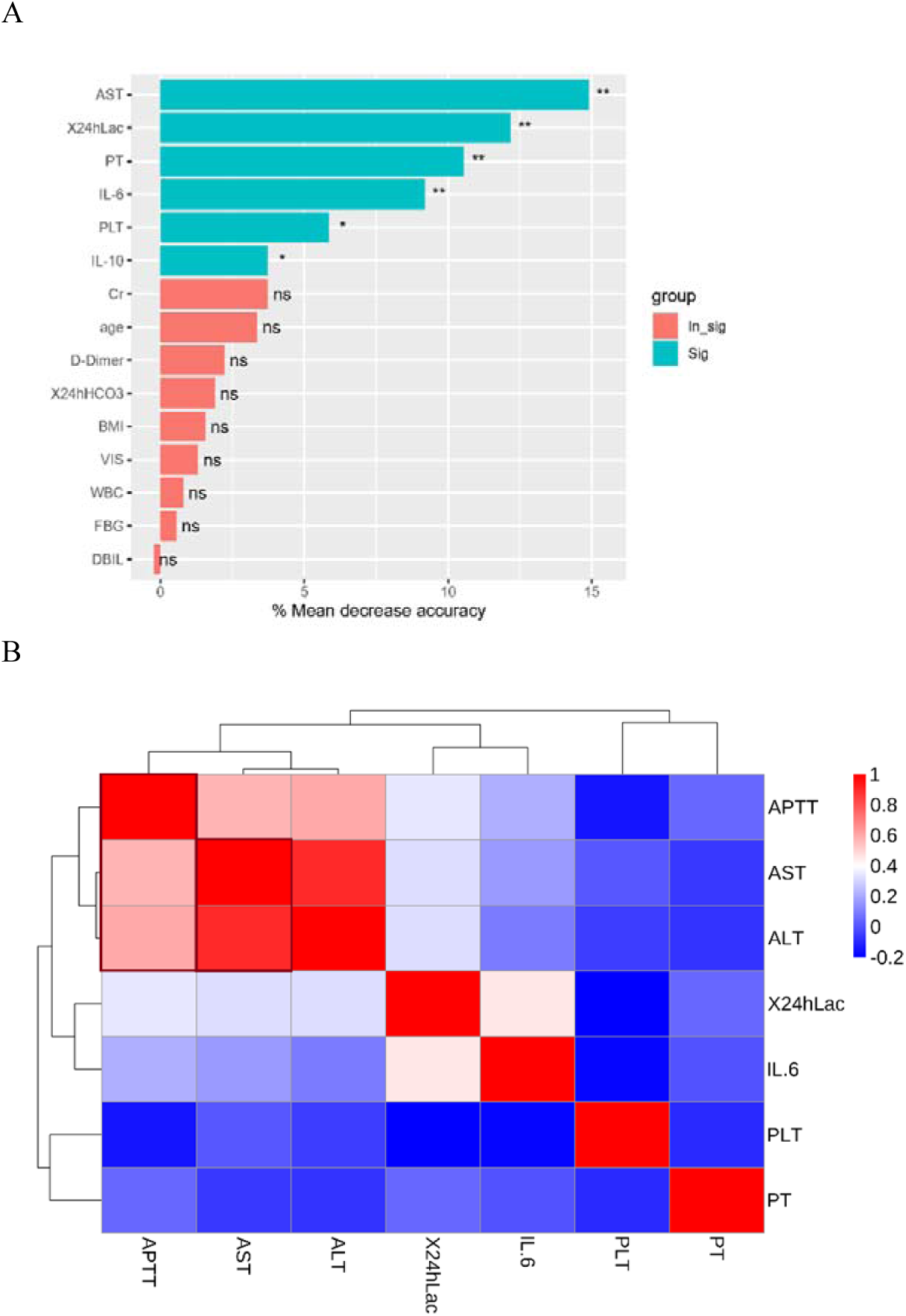
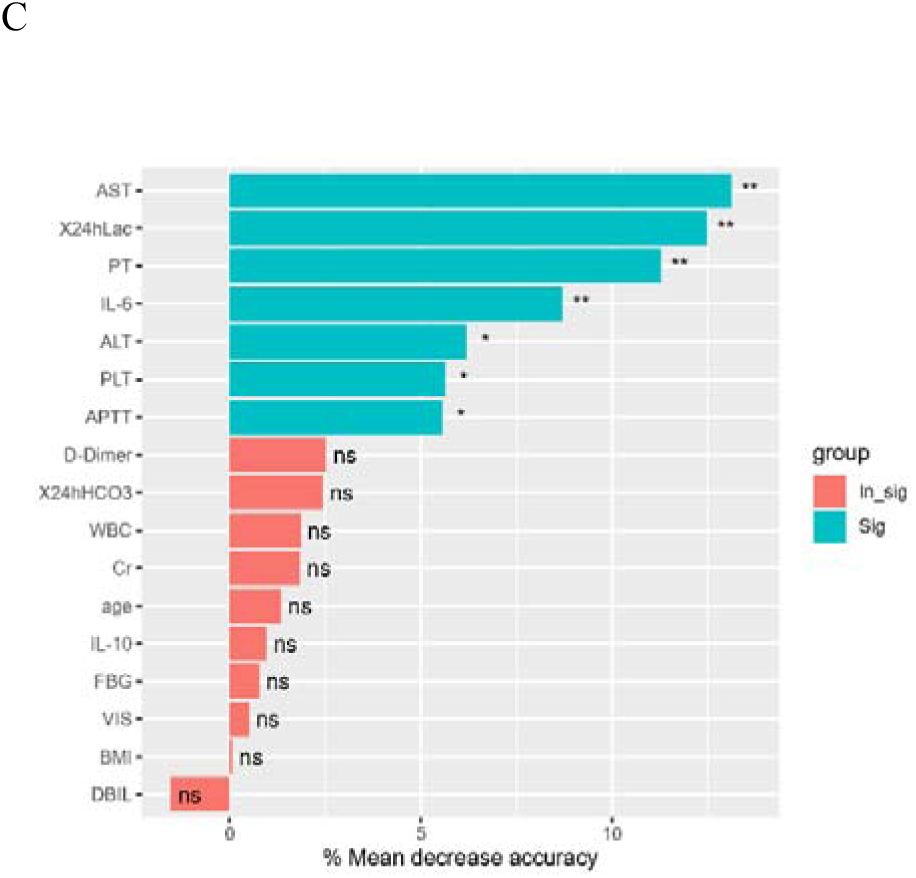
Selection of cluster-determined variables. **A**: A Random Forest Classifier was trained on in-hospital mortality to identify the most mortality-driving variables. Figure 2A shows the result using all variables (including collinear variables). **B**: Out of the most predictive variables, the correlating (collinear) variables were identified using a correlation matrix, and pairs of correlating (|r| > 0.6) variables with a lower predictive value than the respective other variables (i.e., APTT and ALT) were removed. **C**: The five variables with the highest value of predicting in-hospital mortality were the same in both instances (before and after excluding the colinear variables)

Next, consensus k-means clustering algorithm analysis revealed that k = 3 had the highest cluster stability (**Figure 3A**), and different metrics of ML algorithm analysis favored k=3 as the optimal choice for characteristics of the three clusters (**Figure 3B**, **3C**, **3D**). The TSNE plot showed a reduction in the dimensionality of the characteristics of the three clusters, which illustrated the distinct differences among the three clusters (**Figure 3E**).

**Figure 3.**
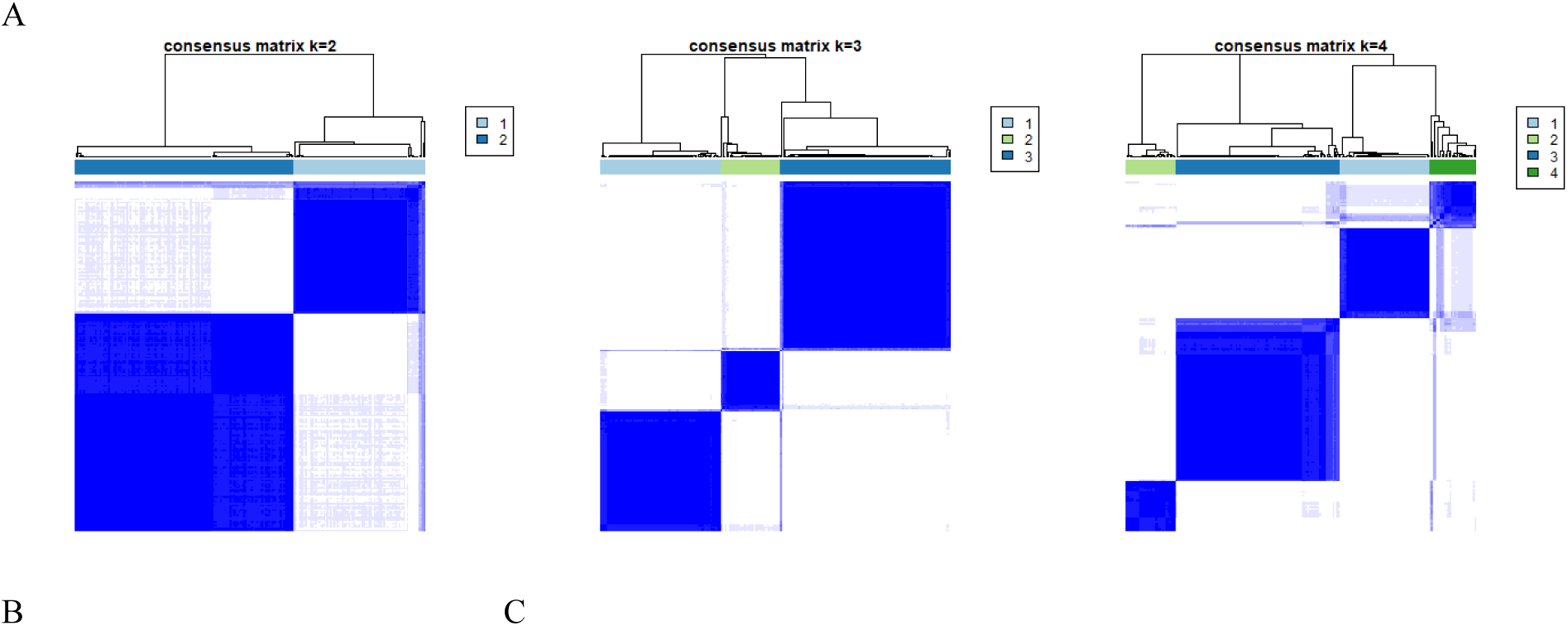
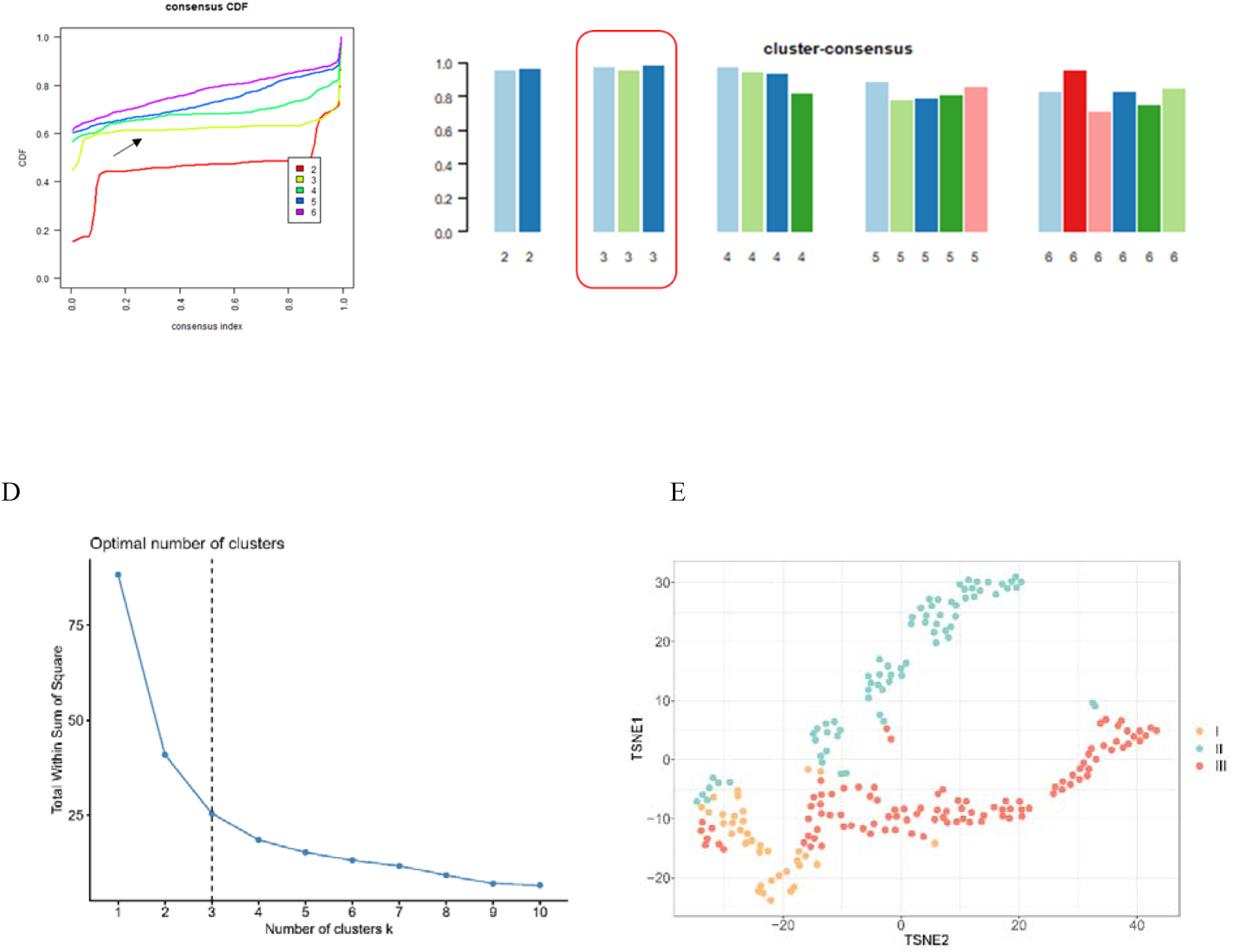
Selecting the number of clusters. A: Comparation of plots graphs with k (number of clusters) = 2, 3, and 4; each column represents one patient, whereas each row displays the assigned clusters. “Sharply marginated” squares indicate stable clusters. K=3 shows the highest cluster stability. B Cumulative Distribution Function (CDF) plot for each k to determine where the CDF reaches a maximum without expense of consensus. Higher a “flatter” curves are favorable (black arrow). C: Cluster-Consensus Plot showing the cluster-consensus values of clusters at each k. High values indicate cluster stability, suggesting that 3 may be the optimal choice for the number of clusters.

Consensus k-means clustering algorithm analysis generated three distinct clusters demonstrating different features, except for the cluster-determined variables. Detailed characteristics, ECMO procedures, laboratory tests, and outcomes are shown in **Table 1**-**4**. In-hospital mortality differed among the clusters, suggesting the effectiveness of the clustering algorithm (**Table 4**). Furthermore, 36 (17.1%), 72 (34.3%), and 102 (48.6%) patients were classified into Cluster I, II, and III, respectively. Radar plots (**Figure 4**) show the deviation of the major laboratory tests (standardized values). According to the clinical profiles, the three most distinctive phenotypes were “platelet preserved (I),” “hyperinflammatory (II),” and “hepatic-renal (III).”

**Figure 4.**
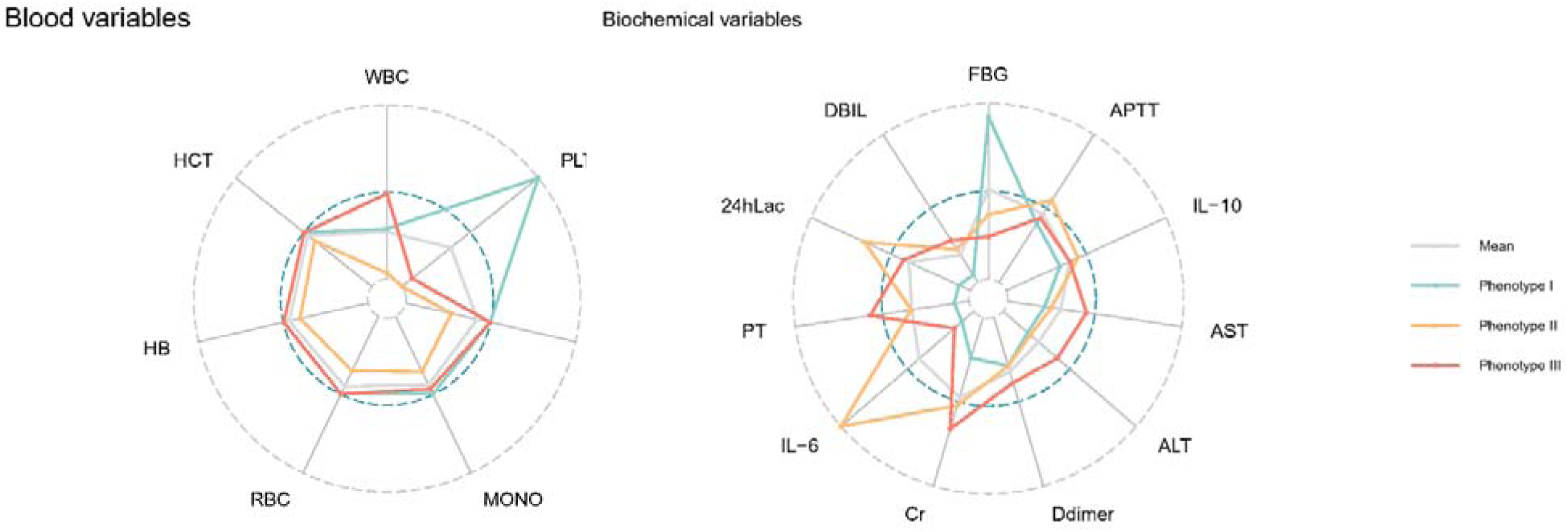
Blood routine and biochemical examination of phenotypes. Radar plots illustrate the characters of blood routine and biochemical examination of each cluster.

### Distinctive features of phenotypes

Phenotype “platelet preserved (I)” had the most preserved quantity of platelet and the highest fibrinogen (FBG) level, the lowest level of the inflammatory-related indicators (IL-6 and IL-10), and preferable liver and kidney function after ECMO initiation (**Table 2**). Therefore, patients seldom needed continuous renal replacement therapy (CRRT) (13.9%) during ECMO support. Compared with the other two phenotypes, fewer patients underwent cardiac procedures (75.0%), especially coronary artery bypass grafting (22.2%) (**Table S1**). This phenotype also had the lowest SOFA scores (**Table 3**) and showed a more sensitive reaction toward ECMO support according to dynamic lactate changes in arterial blood gas examination (**Figure 5**).

**Figure 5.**
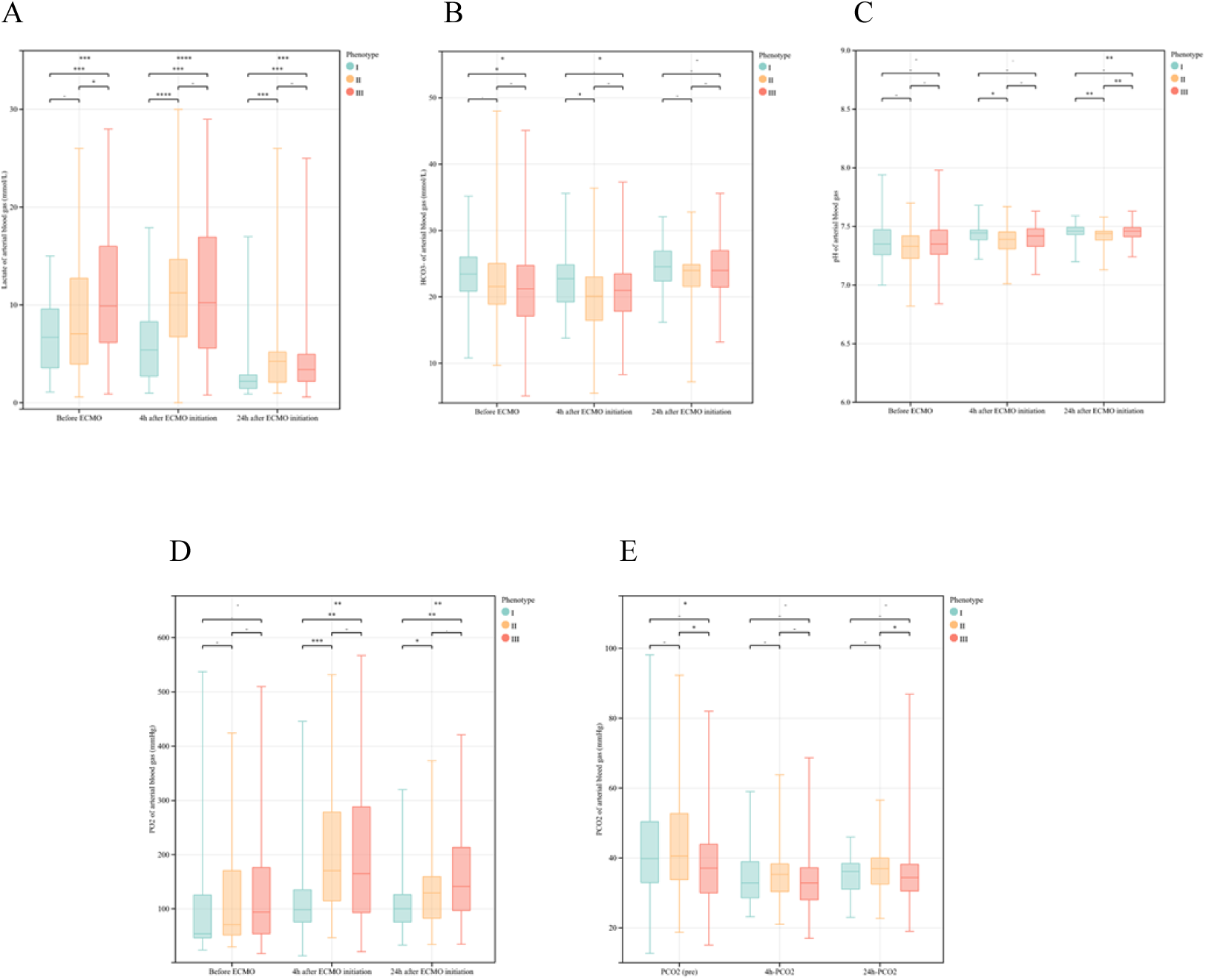
Phenotype reactions to ECMO. A, B, C, D and E show the dynamic changes lactate level, HCO3-level, pH, PO2 and PCO2 among three phenotypes respectively, indicating separated status towards ECMO support.

**Table 2.**
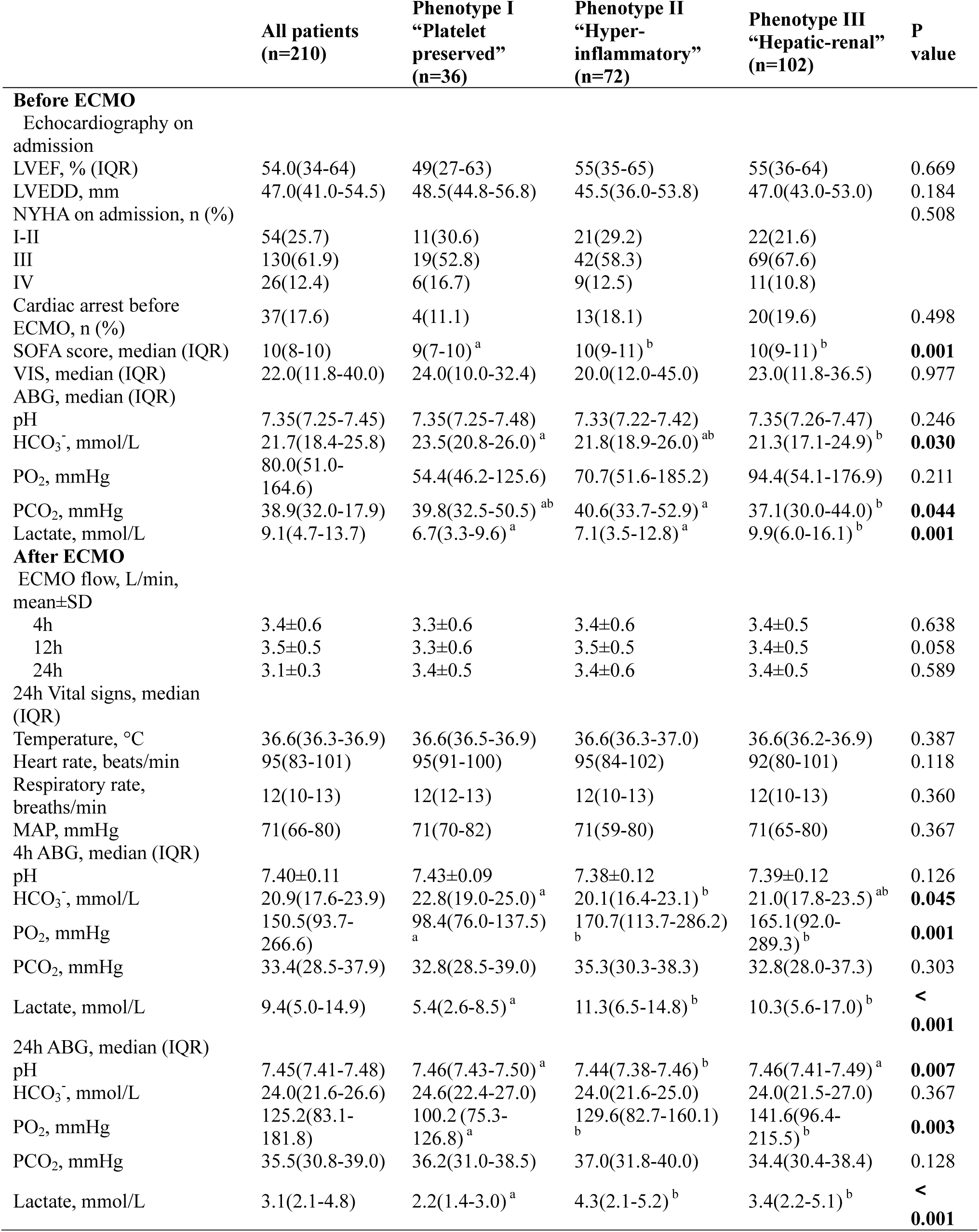
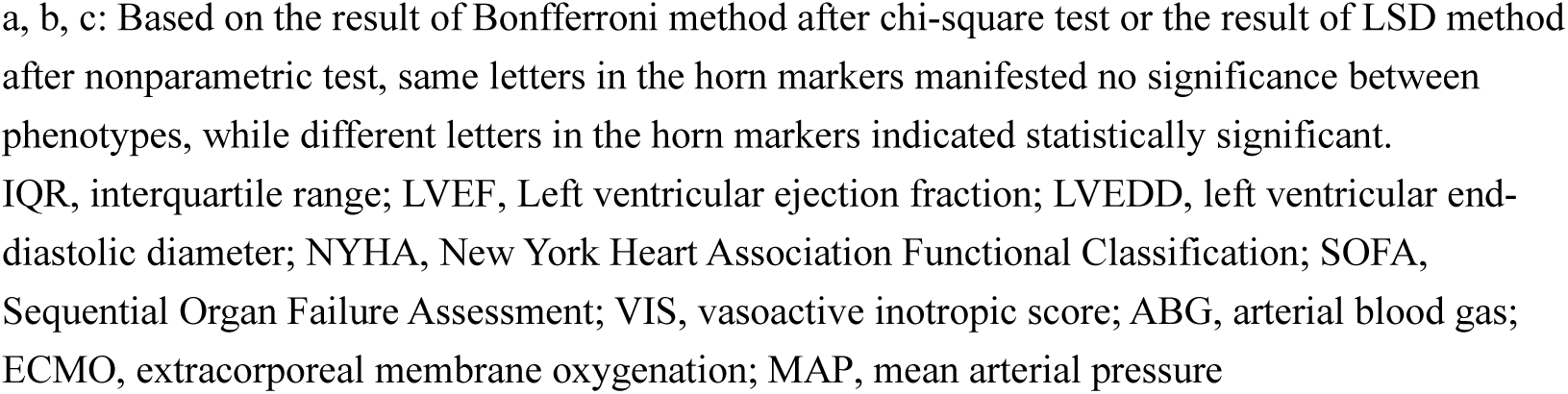
ECMO procedures

|  | All patients<br>(n=210) | Phenotype I<br>“Platelet<br>preserved”<br>(n=36) | Phenotype II<br>“Hyper-<br>inflammatory”<br>(n=72) | Phenotype III<br>“Hepatic-renal”<br>(n=102) | P<br>value |
| --- | --- | --- | --- | --- | --- |
| **Before ECMO** |  |  |  |  |  |
| Echocardiography on admission |  |  |  |  |  |
| LVEF, % (IQR) | 54.0(34-64) | 49(27-63) | 55(35-65) | 55(36-64) | 0.669 |
| LVEDD, mm | 47.0(41.0-54.5) | 48.5(44.8-56.8) | 45.5(36.0-53.8) | 47.0(43.0-53.0) | 0.184 |
| NYHA on admission, n (%) |  |  |  |  | 0.508 |
| I-II | 54(25.7) | 11(30.6) | 21(29.2) | 22(21.6) |  |
| III | 130(61.9) | 19(52.8) | 42(58.3) | 69(67.6) |  |
| IV | 26(12.4) | 6(16.7) | 9(12.5) | 11(10.8) |  |
| Cardiac arrest before ECMO, n (%) | 37(17.6) | 4(11.1) | 13(18.1) | 20(19.6) | 0.498 |
| SOFA score, median (IQR) | 10(8-10) | 9(7-10)<sup>a</sup> | 10(9-11)<sup>b</sup> | 10(9-11)<sup>b</sup> | **0.001** |
| VIS, median (IQR) | 22.0(11.8-40.0) | 24.0(10.0-32.4) | 20.0(12.0-45.0) | 23.0(11.8-36.5) | 0.977 |
| ABG, median (IQR) |  |  |  |  |  |
| pH | 7.35(7.25-7.45) | 7.35(7.25-7.48) | 7.33(7.22-7.42) | 7.35(7.26-7.47) | 0.246 |
| HCO<sub>3</sub><sup>-</sup>, mmol/L | 21.7(18.4-25.8) | 23.5(20.8-26.0)<sup>a</sup> | 21.8(18.9-26.0)<sup>ab</sup> | 21.3(17.1-24.9)<sup>b</sup> | **0.030** |
| PO<sub>2</sub>, mmHg | 80.0(51.0-164.6) | 54.4(46.2-125.6) | 70.7(51.6-185.2) | 94.4(54.1-176.9) | 0.211 |
| PCO<sub>2</sub>, mmHg | 38.9(32.0-17.9) | 39.8(32.5-50.5)<sup>ab</sup> | 40.6(33.7-52.9)<sup>a</sup> | 37.1(30.0-44.0)<sup>b</sup> | **0.044** |
| Lactate, mmol/L | 9.1(4.7-13.7) | 6.7(3.3-9.6)<sup>a</sup> | 7.1(3.5-12.8)<sup>a</sup> | 9.9(6.0-16.1)<sup>b</sup> | **0.001** |
| **After ECMO** |  |  |  |  |  |
| ECMO flow, L/min, mean±SD |  |  |  |  |  |
| 4h | 3.4±0.6 | 3.3±0.6 | 3.4±0.6 | 3.4±0.5 | 0.638 |
| 12h | 3.5±0.5 | 3.3±0.6 | 3.5±0.5 | 3.4±0.5 | 0.058 |
| 24h | 3.1±0.3 | 3.4±0.5 | 3.4±0.6 | 3.4±0.5 | 0.589 |
| 24h Vital signs, median (IQR) |  |  |  |  |  |
| Temperature, °C | 36.6(36.3-36.9) | 36.6(36.5-36.9) | 36.6(36.3-37.0) | 36.6(36.2-36.9) | 0.387 |
| Heart rate, beats/min | 95(83-101) | 95(91-100) | 95(84-102) | 92(80-101) | 0.118 |
| Respiratory rate, breaths/min | 12(10-13) | 12(12-13) | 12(10-13) | 12(10-13) | 0.360 |
| MAP, mmHg | 71(66-80) | 71(70-82) | 71(59-80) | 71(65-80) | 0.367 |
| 4h ABG, median (IQR) |  |  |  |  |  |
| pH | 7.40±0.11 | 7.43±0.09 | 7.38±0.12 | 7.39±0.12 | 0.126 |
| HCO<sub>3</sub><sup>-</sup>, mmol/L | 20.9(17.6-23.9) | 22.8(19.0-25.0)<sup>a</sup> | 20.1(16.4-23.1)<sup>b</sup> | 21.0(17.8-23.5)<sup>ab</sup> | **0.045** |
| PO<sub>2</sub>, mmHg | 150.5(93.7-266.6) | 98.4(76.0-137.5)<sup>a</sup> | 170.7(113.7-286.2)<sup>b</sup> | 165.1(92.0-289.3)<sup>b</sup> | **0.001** |
| PCO<sub>2</sub>, mmHg | 33.4(28.5-37.9) | 32.8(28.5-39.0) | 35.3(30.3-38.3) | 32.8(28.0-37.3) | 0.303 |
| Lactate, mmol/L | 9.4(5.0-14.9) | 5.4(2.6-8.5)<sup>a</sup> | 11.3(6.5-14.8)<sup>b</sup> | 10.3(5.6-17.0)<sup>b</sup> | **< 0.001** |
| 24h ABG, median (IQR) |  |  |  |  |  |
| pH | 7.45(7.41-7.48) | 7.46(7.43-7.50)<sup>a</sup> | 7.44(7.38-7.46)<sup>b</sup> | 7.46(7.41-7.49)<sup>a</sup> | **0.007** |
| HCO<sub>3</sub><sup>-</sup>, mmol/L | 24.0(21.6-26.6) | 24.6(22.4-27.0) | 24.0(21.6-25.0) | 24.0(21.5-27.0) | 0.367 |
| PO<sub>2</sub>, mmHg | 125.2(83.1-181.8) | 100.2(75.3-126.8)<sup>a</sup> | 129.6(82.7-160.1)<sup>b</sup> | 141.6(96.4-215.5)<sup>b</sup> | **0.003** |
| PCO<sub>2</sub>, mmHg | 35.5(30.8-39.0) | 36.2(31.0-38.5) | 37.0(31.8-40.0) | 34.4(30.4-38.4) | 0.128 |
| Lactate, mmol/L | 3.1(2.1-4.8) | 2.2(1.4-3.0)<sup>a</sup> | 4.3(2.1-5.2)<sup>b</sup> | 3.4(2.2-5.1)<sup>b</sup> | **< 0.001** |

a, b, c: Based on the result of Bonferroni method after chi-square test or the result of LSD method after nonparametric test, same letters in the horn markers manifested no significance between phenotypes, while different letters in the horn markers indicated statistically significant.
IQR, interquartile range; LVEF, Left ventricular ejection fraction; LVEDD, left ventricular end-diastolic diameter; NYHA, New York Heart Association Functional Classification; SOFA, Sequential Organ Failure Assessment; VIS, vasoactive inotropic score; ABG, arterial blood gas; ECMO, extracorporeal membrane oxygenation; MAP, mean arterial pressure

Phenotype “hyperinflammatory (II)” mainly consisted of male patients manifesting a statistically significant increase in inflammatory indicators, such as IL-6 and IL-10 (**Table 3, Figure 4**). The APTT of this phenotype was significantly prolonged compared with that of the others, and the liver function was worse than that of phenotype I but better than that of phenotype III (**Table 3**). In comparison, renal function was in the same poor condition as that of phenotype III. There was no significant difference in the reaction towards ECMO support compared with that of phenotype III (**Table 2**, **Figure 5**).

**Table 3.**
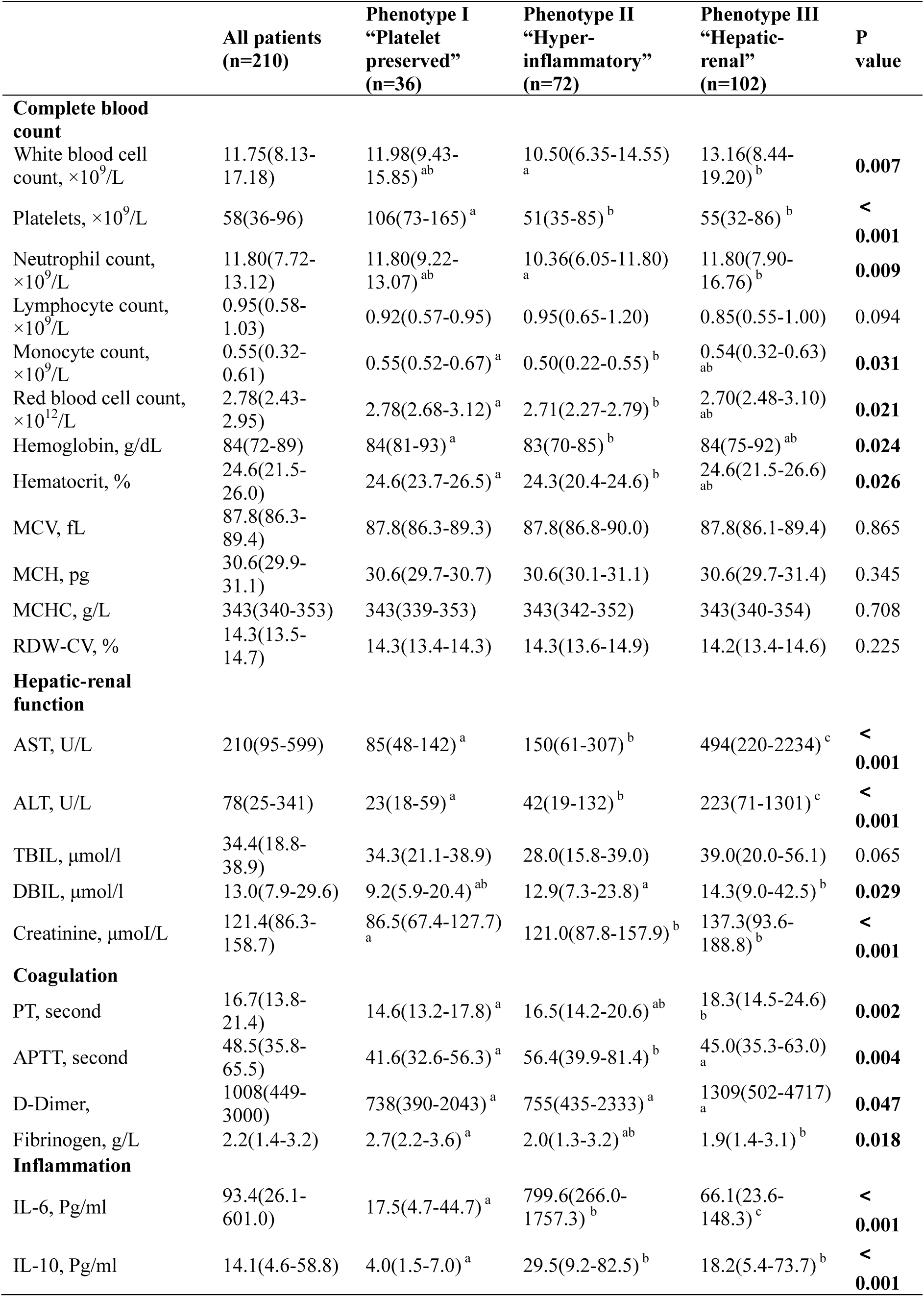
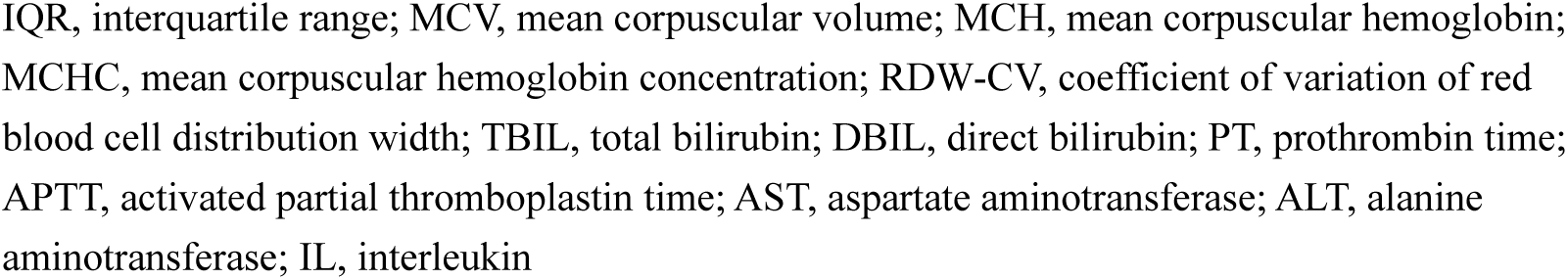
Laboratory tests of 24 hours after ECMO initiation

|  | All patients<br>(n=210) | Phenotype I<br>“Platelet<br>preserved”<br>(n=36) | Phenotype II<br>“Hyper-<br>inflammatory”<br>(n=72) | Phenotype III<br>“Hepatic-renal”<br>(n=102) | P<br>value |
| --- | --- | --- | --- | --- | --- |
| **Complete blood count** |  |  |  |  |  |
| White blood cell count, $\times 10^9/L$ | 11.75(8.13-17.18) | 11.98(9.43-15.85)<sup>ab</sup> | 10.50(6.35-14.55)<sup>a</sup> | 13.16(8.44-19.20)<sup>b</sup> | **0.007** |
| Platelets, $\times 10^9/L$ | 58(36-96) | 106(73-165)<sup>a</sup> | 51(35-85)<sup>b</sup> | 55(32-86)<sup>b</sup> | **< 0.001** |
| Neutrophil count, $\times 10^9/L$ | 11.80(7.72-13.12) | 11.80(9.22-13.07)<sup>ab</sup> | 10.36(6.05-11.80)<sup>a</sup> | 11.80(7.90-16.76)<sup>b</sup> | **0.009** |
| Lymphocyte count, $\times 10^9/L$ | 0.95(0.58-1.03) | 0.92(0.57-0.95) | 0.95(0.65-1.20) | 0.85(0.55-1.00) | 0.094 |
| Monocyte count, $\times 10^9/L$ | 0.55(0.32-0.61) | 0.55(0.52-0.67)<sup>a</sup> | 0.50(0.22-0.55)<sup>b</sup> | 0.54(0.32-0.63)<sup>ab</sup> | **0.031** |
| Red blood cell count, $\times 10^{12}/L$ | 2.78(2.43-2.95) | 2.78(2.68-3.12)<sup>a</sup> | 2.71(2.27-2.79)<sup>b</sup> | 2.70(2.48-3.10)<sup>ab</sup> | **0.021** |
| Hemoglobin, g/dL | 84(72-89) | 84(81-93)<sup>a</sup> | 83(70-85)<sup>b</sup> | 84(75-92)<sup>ab</sup> | **0.024** |
| Hematocrit, % | 24.6(21.5-26.0) | 24.6(23.7-26.5)<sup>a</sup> | 24.3(20.4-24.6)<sup>b</sup> | 24.6(21.5-26.6)<sup>ab</sup> | **0.026** |
| MCV, fL | 87.8(86.3-89.4) | 87.8(86.3-89.3) | 87.8(86.8-90.0) | 87.8(86.1-89.4) | 0.865 |
| MCH, pg | 30.6(29.9-31.1) | 30.6(29.7-30.7) | 30.6(30.1-31.1) | 30.6(29.7-31.4) | 0.345 |
| MCHC, g/L | 343(340-353) | 343(339-353) | 343(342-352) | 343(340-354) | 0.708 |
| RDW-CV, % | 14.3(13.5-14.7) | 14.3(13.4-14.3) | 14.3(13.6-14.9) | 14.2(13.4-14.6) | 0.225 |
| **Hepatic-renal function** |  |  |  |  |  |
| AST, U/L | 210(95-599) | 85(48-142)<sup>a</sup> | 150(61-307)<sup>b</sup> | 494(220-2234)<sup>c</sup> | **< 0.001** |
| ALT, U/L | 78(25-341) | 23(18-59)<sup>a</sup> | 42(19-132)<sup>b</sup> | 223(71-1301)<sup>c</sup> | **< 0.001** |
| TBIL, $\mu\text{mol/l}$ | 34.4(18.8-38.9) | 34.3(21.1-38.9) | 28.0(15.8-39.0) | 39.0(20.0-56.1) | 0.065 |
| DBIL, $\mu\text{mol/l}$ | 13.0(7.9-29.6) | 9.2(5.9-20.4)<sup>ab</sup> | 12.9(7.3-23.8)<sup>a</sup> | 14.3(9.0-42.5)<sup>b</sup> | **0.029** |
| Creatinine, $\mu\text{mol/L}$ | 121.4(86.3-158.7) | 86.5(67.4-127.7)<sup>a</sup> | 121.0(87.8-157.9)<sup>b</sup> | 137.3(93.6-188.8)<sup>b</sup> | **< 0.001** |
| **Coagulation** |  |  |  |  |  |
| PT, second | 16.7(13.8-21.4) | 14.6(13.2-17.8)<sup>a</sup> | 16.5(14.2-20.6)<sup>ab</sup> | 18.3(14.5-24.6)<sup>b</sup> | **0.002** |
| APTT, second | 48.5(35.8-65.5) | 41.6(32.6-56.3)<sup>a</sup> | 56.4(39.9-81.4)<sup>b</sup> | 45.0(35.3-63.0)<sup>a</sup> | **0.004** |
| D-Dimer, | 1008(449-3000) | 738(390-2043)<sup>a</sup> | 755(435-2333)<sup>a</sup> | 1309(502-4717)<sup>a</sup> | **0.047** |
| Fibrinogen, g/L | 2.2(1.4-3.2) | 2.7(2.2-3.6)<sup>a</sup> | 2.0(1.3-3.2)<sup>ab</sup> | 1.9(1.4-3.1)<sup>b</sup> | **0.018** |
| **Inflammation** |  |  |  |  |  |
| IL-6, Pg/ml | 93.4(26.1-601.0) | 17.5(4.7-44.7)<sup>a</sup> | 799.6(266.0-1757.3)<sup>b</sup> | 66.1(23.6-148.3)<sup>c</sup> | **< 0.001** |
| IL-10, Pg/ml | 14.1(4.6-58.8) | 4.0(1.5-7.0)<sup>a</sup> | 29.5(9.2-82.5)<sup>b</sup> | 18.2(5.4-73.7)<sup>b</sup> | **< 0.001** |

IQR, interquartile range; MCV, mean corpuscular volume; MCH, mean corpuscular hemoglobin; MCHC, mean corpuscular hemoglobin concentration; RDW-CV, coefficient of variation of red blood cell distribution width; TBIL, total bilirubin; DBIL, direct bilirubin; PT, prothrombin time; APTT, activated partial thromboplastin time; AST, aspartate aminotransferase; ALT, alanine aminotransferase; IL, interleukin

Phenotype “hepatic-renal (III)” had poor liver function (elevated AST, ALT, PT, DBIL, and FBG levels) (**Table 3**) and the highest serum creatinine level among the clusters. The sensitivity of the reaction towards ECMO support (24 h after ECMO initiation) was significantly poor in terms of elimination of lactate and oxygen consumption (**Figure 5**). The tendencies of the median standardized values of the cluster-determined variables are shown in **Figure 6**.

**Figure 6.**
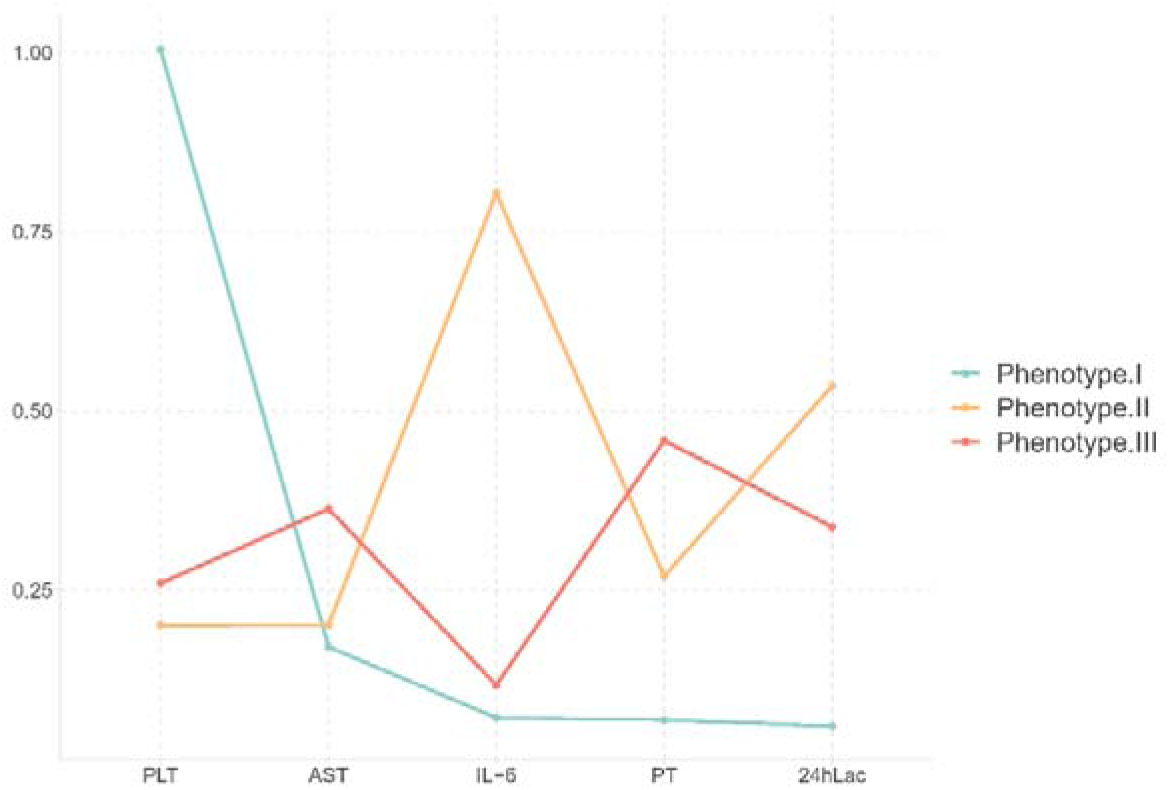
Tendency of Median Standardized Values of Cluster-determined Variables. Line charts showed distinct tendency of cluster-determined variables among clusters PLT, Platelet count; AST’ Aspartic acid transaminase; IL-6, Interleukin-6; PT, Prothrombin time; ECMO24h-Lac, Lactate level of arterial blood at 24 hours after ECMO initiation.

### Phenotype reactions to ECMO

Dynamic changes in arterial blood gas revealed different reaction tendencies toward 24-hour ECMO support among the three phenotypes (**Table 2**, **Figure 5**). Significant changes occurred in the lactate and oxygen partial pressure (PaO_2_) of arterial blood. Before ECMO implantation, the lactate level of Phenotype III was the highest among clusters, and after 4 h and 24 h of ECMO support, the lactate level of Phenotype I was significantly lower than that of the others. A similar trend also existed in the change of PO_2_. There was no statistical difference among phenotypes before ECMO initiation, but significant changes occurred after 4 h and 24 h of ECMO support (patients in Phenotype I gradually showed a significantly lower level of PO2 than the others). Other details of ECMO, such as peak blood flow and vital signs, did not show any significant differences (**Table 3**).

### Outcomes

The in-hospital mortality rates of phenotypes I, II, and III were 25.0%, 52.8%, and 55.9%, respectively. Compared to Phenotype I, patients with Phenotype II had higher mortality (odds ratio [OR], 2.3 [95% CI, 1.2–4.4]), whereas those with Phenotype III (OR, 2.8 [95% CI, 1.4–5.4]) had the highest mortality. There were no significant differences in the other outcomes among phenotypes, such as length of hospital and ICU stay, bleeding complications, and limb ischemia (**Table 4**).

**Table 4.** Outcomes

Neurological complications including cerebral hemorrhage, cerebral infarction, epileptic seizure,
|  | All patients (n=210) | Phenotype I<br>“Platelet<br>preserved”<br>(n=36) | Phenotype II<br>“Hyper-<br>inflammatory”<br>(n=72) | Phenotype III<br>“Hepatic-renal”<br>(n=102) | P value |
| --- | --- | --- | --- | --- | --- |
| In-hospital mortality | 104(49.5) | 9(25.0)<sup>a</sup> | 38(52.8)<sup>b</sup> | 57(55.9)<sup>b</sup> | **0.005** |
| Successful weaning from ECMO | 142(67.6) | 30(83.3) | 45(62.5) | 67(65.7) | 0.078 |
| ECMO duration (hour) | 105.4(66.7-153.6) | 98.5(87.4-132.0) | 105.0(48.9-147.9) | 108.0(68-183) | 0.310 |
| MV duration (hour) | 190.7(82.4-269.9) | 193.5(99.9-302.1) | 105.4(66.6-153.6) | 150.7(90.5-285.2) | 0.547 |
| Complications |  |  |  |  |  |
| Bleeding at intubation site | 25(13) | 2(6.1) | 8(11.9) | 15(16.1) | 0.319 |
| Limb ischemia required fasciotomy | 9(4.3) | 1(2.8) | 4(5.6) | 4(1.9) | 0.773 |
| Cr > 3.0 mg/dL | 32(15.2) | 1(2.8) | 11(15.3) | 20(19.6) | 0.054 |
| Hyperbilirubinemia | 104(49.5) | 5(13.9)<sup>a</sup> | 43(59.7)<sup>b</sup> | 56(54.9)<sup>b</sup> | **< 0.001** |
| Neurological complications | 15(7.1) | 3(8.3) | 7(9.7) | 5(4.9) | 0.456 |
| Hospital length of stay (day) | 20(12-28) | 20(15-30) | 16(10-26) | 21(13-29) | 0.096 |
| ICU length of stay (day) | 9(5-14) | 10(6-14) | 9(3-13) | 10(4-15) | 0.231 |

and cerebral death.
Hyperbilirubinemia was defined as DBIL > 2 mg/dL, IBIL > 13 mg/dL, or TBIL > 15 mg/dL.
IQR, interquartile range; ICU, intensive care unit; ECMO, extracorporeal membrane oxygenation

## Discussion

In the cohort of refractory patients with CS receiving VA-ECMO, we used consensus k-means algorithm analysis, a classical machine learning algorithm, to profile the potential heterogeneity of these disease statuses for the first time. After strict clustering number determination and verification, we identified three clusters of patients with various clinical manifestations, inflammatory profiles, and outcomes. The three clusters were noted as the “platelet preserved (I),” “hyperinflammatory (II),” and “hepatic-renal (III)” phenotypes. Accordingly, these findings may improve risk stratification, enable the development of treatment algorithms tailored to each phenotype of patients with ECMO, and inform patient selection in future clinical trials.

The CS patients in this cohort were mostly men diagnosed with coronary artery disease, and the majority of them underwent cardiac procedures. The in-hospital mortality rate of this cohort was 49.5%, which was consistent with a previous report of 43%–60% [25–29]. The simplification of VA-ECMO implantation and the gradually matured strategy benefited an increasing number of patients with CS, regardless of the treatment period or clinical outcomes. However, patients with CS receiving ECMO still had considerable variability in terms of in-hospital mortality. Therefore, much effort has been put into determining prognosis-related factors or prediction models of ECMO patient outcomes. However, the heterogeneity of disease status may profoundly affect the prognosis of the disease, as in patients with CS, which means that the single dimension of predicting strategy performs poorly.

New approaches, such as k-means algorithm analysis and latent class analysis (LCA), have been implemented to define the distinct clinical phenotypes of diseases. For instance, the heterogeneity of acute respiratory distress syndrome (ARDS) was reported by Calfee et al. They used clinical and biological data from two ARDS randomized controlled trials and applied latent class modeling to identify two distinct phenotypes [30]. ARDS phenotypes were later confirmed by studies performed on randomized controlled trial (RCT) cohorts. Patients with the two subtypes appear to have different benefits in distinct fluid and pharmacotherapeutic strategies [12, 31]. These heterogeneities have also been widely studied during the coronavirus epidemic [32, 33]. In the present study, we analyzed the clinical presentations, sensitivity to ECMO support, prognosis, and inflammatory profiles in detail after cluster analysis to generalize the heterogeneous characteristics of distinct phenotypes.

In this VA-ECMO cohort, the “Platelet preserved (I)” phenotype represented a preserved platelet count, correlating with a favorable prognosis. Thrombocytopenia and platelet dysfunction are common in patients with ECMO, regardless of the ECMO mode. It has been demonstrated that more than 20% have platelet counts lower than 150 × 10^9^/L at some points during VA-ECMO [9]. External circuit surfaces and high shear stress during ECMO treatment play major roles in platelet activation and aggregation [34, 35]. Thrombocytopenia develops after cardiac surgery and ECMO because of extensive crosstalk between inflammation, coagulation, bleeding, extracorporeal circuit consumption, and oxidizing stress caused by high oxygen tension [8, 36]. Thrombocytopenia has also been proven to be an independent risk factor for poor outcomes in patients undergoing ECMO after cardiac surgery [8, 37]. Persistent, severe thrombocytopenia indicates a significant physiologic imbalance [37]. Inversely, a higher platelet count may indicate a better immune and pathophysiologic state. This is consistent with our study. Phenotype I had a mild inflammatory response and a lower IL-6 level.

As for the reaction towards ECMO after 24 h of support, phenotype I patients had no significant difference from the other clusters in lactate level in arterial blood before ECMO implantation. However, the difference occurred with prolonged treatment, and the level of lactate in phenotype I was the lowest among clusters after both 4 h and 24 h (**Table 2**, **Figure 5**). Lactate behavior is a classic and vital factor related to in-hospital mortality, and has been widely discussed. Several studies have highlighted the independently predicted value of survival of patients with CS [6, 7, 38, 39]. The lactate scale (<2, 2-8, or >8 mmol/L) has even been identified as an independent predictor of mortality in patients undergoing VA-ECMO after coronary artery bypass grafting by scoring systems such as the REMEMBER score [40]. The pathophysiologic status generalized in phenotype I indicates that respiratory and circulatory function, as well as tissue perfusion of these patients, recovered promptly, manifesting a gradual decrease in PO_2_ and a lower level of AST and ALT. This was the direct opposite of the pathophysiological status in phenotype III.

Similar to the lactate level, there were no statistical differences in the PaO_2_ of arterial blood among clusters before ECMO initiation. However, significant differences occurred 4 h and 24 h after ECMO implantation, “platelet preserved (I)” phenotype patients displayed the lowest level of PaO2 of arterial blood compared with the others. Hyperoxia increases oxidative stress, producing free radicals and reactive oxygen species (ROS), which promote neutrophil activation and lead to an inappropriate inflammatory response. This effect can be amplified by the complex status of critically ill patients with mechanical circulatory assistance [41]. Clinical reports on oxygen management during VA-ECMO support for CS are limited. Recent research manifested a significant association between hyperoxia and mortality during ECMO [25, 42]. Moreover, Moussa et al. found that even a very short exposure to hyperoxia was harmless for patients after receiving ECMO support [25], which is consistent with our finding that patients with a lower level of PO_2_ in the first 24 h after ECMO initiation correlated with a favorable prognosis.

Our findings also revealed the existence of a hyperinflammatory subtype (phenotype II) in ECMO patients, which correlated with an unfavorable prognosis. It was widely approved that inflammatory conditions and oxidative stress could affect the outcome of patients receiving VA-ECMO, as evidenced by significant induction of various inflammatory mediators (such as various interleukins (ILs) including IL-1β, IL-6, IL-8, IL-18, and TNF-α) and markers of oxidative stress (such as oxidized low-density lipoprotein (ox-LDL), and malondialdehyde (MDA)) [19, 43]. We detected IL-6 and IL-8 as the representative cytokines to profile the inflammatory status in ML approaches, and found a distinct tendency of cytokine values among clusters. This was also the first study to use inflammatory mediators to perform the ML algorithm on patients with CS receiving ECMO support. Emerging observational data have demonstrated the efficacy of cytokine adsorption in patients supported by ECMO [43]. The hyperinflammatory phenotype might provide selected patient candidates for further trials of cytokine adsorption to diminish the inflammatory response during ECMO support.

Patients with phenotype III were characterized by worsening renal and hepatic function, which appeared to develop into multiorgan dysfunction and refractory phase with the highest in-hospital mortality. This was consistent with a previous study on cardiogenic shock using the clustering algorithm, which also obtained a cluster with hepatorenal function disorder and a high mortality rate [14]. “Organ crosstalk,” which refers to bidirectional interactions between distant organs, summarizes the complex biological communication and feedback that occurs between different organs mediated via numerous mechanisms [44]. Renal function and congestion have been identified as important prognostic factors for the outcomes in patients with acute and chronic heart failure [45]. Previous reports found that more than 70% of patients receiving ECMO developed acute kidney injury (AKI), and AKI requiring renal replacement therapy (RRT) in patients undergoing ECMO treatment increased mortality in ICU patients [46, 47]. The liver plays a vital role in oxidant scavenging and antioxidative replenishment in the body, and may be more susceptible to inflammation and oxidative stress during extracorporeal circulation [48]. In phenotype III, corrupted hepatic-renal function reflects a refractory tissue perfusion disorder, leading to multiple organ disorder syndrome (MODS) in the absence of immediate treatment. Therefore, the timing and strategy of RRT or multiple organ support are of great importance for patients with phenotype III.

Although the risk of death was the highest in phenotype III, there was no significant difference between phenotypes II and III. The interaction between inflammatory and oxidative stress and organ function suggests that there may exist an overlapping trend between phenotypes II and III. Before developing into phenotype III, phenotype II may be the appropriate time for ECMO implantation, which would be beneficial for the patients.

In brief, identifying these distinct phenotypes may provide a novel perspective on furthering the perception of this disease status for clinical staff and may also provide some advice on the development of treatment strategies in subsets of patients with CS receiving ECMO. For instance, clinicians could flexibly change the treatment strategy when a poor tendency is recognized. Moreover, the verification of these findings requires further cohort studies.

### Limitation

First, this was a single-center prospective study, and the main limitation was the small sample size. Logic cluster analysis demanded as many data points as possible to stabilize the prediction model; therefore, it was difficult to build a validation cohort using this restricted scale of cases. Second, small-scale databases limit the diversity of cluster-determined variables, which may mask other underlying traits between subtypes to a certain extent. Third, the validation of the clustering model and the illustration of other potential dimensionalities of cluster features require a larger scale of multi-center data. Lastly, our cohort was mainly composed of postcardiotomy patients, which may cause bias in the distribution of etiology and may amplify the inflammatory response of patients induced by surgery when compared with that of other ECMO cohorts.

## Conclusion

With the consensus k-means algorithm analysis identified three phenotypes in refractory patients with CS receiving VA-ECMO: “platelet preserved,” “hyperinflammatory,” and “hepatic-renal.” The phenotypes are specifically associated with clinical profile and mortality, and they may allow for the development of treatment strategies for subsets of patients with CS receiving ECMO.

## Data Availability

All data referred to in the manuscript is objective and true,and will be available to reviewers if necessary.

## Abbreviations

VA-ECMO: Venoarterial extracorporeal membrane oxygenation
CS: cardiogenic shock
PLT: platelet count
AST: aspartic acid transaminase
IL-6: Interleukin-6
PT: prothrombin time
ML: Machine learning
SAVE: The survival after VA-ECMO score
AMI: acute myocardial infarction
REMEMBER: pRedicting mortality in patients undergoing veno-arterial Extracorporeal MEMBrane oxygenation after coronary artEry bypass gRafting
ENCOURAGE: prEdictioN of Cardiogenic shock OUtcome foR Acute myocardial infarction patients salvaGed by VA-ECMO score
ARDS: acute respiratory distress syndrome
LVEF: left ventricular ejection fraction
LVEDD: left ventricular end-diastolic diameter
NYHA: New York Heart Association Functional Classification
VIS: the vasoactive inotropic score
ABG: arterial blood gas
MAP: mean arterial pressure
TSNE plot: t-distributed stochastic neighbor embedding plot
IQR: Interquartile range
CRRT: Continuous renal replacement therapy
SOFA: The Sepsis-related Organ Failure Assessment score
ROS: reactive oxygen species
ox-LDL: oxidized low-density lipoprotein
MDA: malondialdehyde
MODS: multiple organ disorder syndrome
IABP: intra-aortic balloon pump
CPB: cardiopulmonary bypass
CABG: Coronary artery bypass grafting
ICU: Intensive care unit

## Acknowledgements

We thank Hui Zeng, Chen Chen and Junyan Han for their assistance in the laboratory test of Interleukin-6 and Interleukin-10 used in this manuscript.

## Funding

National Natural Science Foundation of China (No. 82170400 to Xiaotong Hou): The mechanism of erythroid precursors regulate negatively monocytes immune function through Galectin-9-TIM3 pathway.

National Natural Science Foundation of China (No. 81870305 to Xiaotong Hou): The negative regulation mechanism of co-inhibitory molecule TIGIT on the function of inflammatory monocytes in patients supported with ECMO.

Beijing Hospitals Authority Clinical Medicine Development of Special Funding Support, Code: ZYLX202111.

Beijing Hospitals Authority “Ascent Plan”, Code: FDL20190601.

Beijing Key Specialist Project for Major Epidemic Prevention and Control Supported by the National Key Research and Development Program of China (No. 2021YFC2701703)

Beijing Nova Program (No. 2022064)

Social Development Science and Technology Project (CYSF2215) from Chaoyang District Bureau of Science and Technology and Information Technology

## Availability of data and materials

The datasets used and/or analyzed during the current study are available from the corresponding author on reasonable request.

## Authors’ contributions

Shuo Wang, Liangshan Wang, Zhongtao Du, Feng Yang, Xing Hao, Xiaomeng Wang and Chenchen Shao collected and analyzed the patient data. Shuo Wang, Liangshan Wang and Chenglong Li performed the statistical analysis and were major contributors in writing the manuscript. Chenglong, Hong Wang and Xiaotong Hou were responsible for the funds and were participating in the supervision of project design and implementation. All authors read and approved the final manuscript.

## Ethics approval and consent to participate

The study was approved by the institutional ethics committee/review board of the Beijing Anzhen Hospital. Informed consent for demographic, physiological and hospital-outcome data analyses was not obtained because this observational study did not modify existing diagnostic or therapeutic strategies. However, patients and/or relatives were informed about the anonymous data collection and that they could decline inclusion.

## Competing interests

The authors declare that they have no competing interests.

## Declaration

All authors approved the final manuscript and the submission to this journal.

---

**Table S1.** Details of patients receiving cardiac surgery

|  | All patients<br>(n=210) | Phenotype I<br>“Platelet<br>preserved”<br>(n=36) | Phenotype II<br>“Hyper-<br>inflammatory<br>”<br>(n=72) | Phenotype<br>III<br>“Hepatic-renal”<br>(n=102) | P value |
| --- | --- | --- | --- | --- | --- |
| Surgery in this hospitalization, n (%) | 184(87.6) | 27(75.0)<sup>a</sup> | 70(97.2)<sup>b</sup> | 87(85.3)<sup>a</sup> | **0.003** |
| Surgery under CPB, n (%) | 144(68.6) | 22(61.1) | 57(79.2) | 65(63.7) | 0.055 |
| Type of surgery, n (%) |  |  |  |  |  |
| CABG | 71(33.8) | 8(22.2)<sup>a</sup> | 32(44.4)<sup>b</sup> | 31(30.4)<sup>b</sup> | **0.040** |
| Valve procedure | 61(29.0) | 11(30.6) | 23(31.9) | 27(26.5) | 0.708 |
| CABG + valve procedure | 26(12.4) | 3(8.3) | 9(12.5) | 14(13.7) | 0.700 |
| Repair of acute aortic dissection | 12(5.7) | 1(2.8) | 5(6.9) | 6(5.9) | 0.732 |
| Repair of acute aortic dissection + CABG | 7(3.3) | 0(0) | 2(2.8) | 5(4.9) | 0.534 |
| Pulmonary embolectomy | 6(2.9) | 1(2.8) | 2(2.8) | 3(2.9) | 1.000 |
| Heart transplantation | 9(4.3) | 3(8.3) | 1(1.4) | 5(4.9) | 0.169 |
| Others | 11(5.2) | 1(2.8) | 5(6.9) | 5(4.9) | 0.770 |

a, b, c: Based on the result of the Bonferroni method after chi-square test or the result of LSD
method after nonparametric test, the same letters in the horn markers indicate no significance
between clusters, while different letters in the horn markers indicate statistically significant.
CPB, cardiopulmonary bypass; CABG, coronary artery bypass graft

## Notes

### Competing Interest Statement

The authors have declared no competing interest.

